# Geohelminth Infections Among School Children in Qua’an-Pan Local Government Area, Plateau State

**DOI:** 10.1101/2025.04.04.25325233

**Authors:** Waalmoep Gerald Damar, Morgak James Gonap, Gudzan Sow, Wandayi Emmanuel Amlabu

**Affiliations:** Department of Zoology, Ahmadu Bello University, Zaria. Nigeria. Residential address; Opposite ITF office Bukuru Expressway, Jos South Local Government Area, Plateau State, Nigeria; Department of Zoology, Ahmadu Bello University, Zaria. Nigeria

**Keywords:** Geohelminths, Children, Qua’an-Pan, Prevalence

## Abstract

Parasitic infections are crucial public health challenges that are known to affect the wellbeing of primary school children in developing countries such as Nigeria. This study was carried out to assess the current prevalence of geohelminth infections among primary school children in Qua’an-Pan Local Government Area. The study was carried out in five districts and a total of 806 children were randomly selected. Stool samples were collected from each pupil to screen for parasites. Percentages were used to obtain prevalence while chi-square test was used to determine significant difference among variables. Formol-ether concentration technique was used to prepare samples for parasitological examination. The overall prevalence of geohelminth infections was 16.38%. Hookworm, *Ascaris lumbricoides* and *Trichuris trichiura* had prevalence of 12.53%, 2.23% and 1.61% respectively. Qua’an-Pan Local Government Area is endemic for geohelminth infections and the prevalence of geohelminthiasis was not associated with sex, age group, location and school type.

## Introduction

Geohelminths or Soil Transmitted Helminths are parasitic intestinal nematodes whose immature stages require a period of development in the soil. They include *Ascaris lumbricoides* (round worm), *Necator americanus* and *Ancylostoma duodenale* (hookworms), *Trichuris trichiura* (whipworm) and *Strongyloides stercoralis* commonly known as the threadworm.(1,2) Depending on the nematode species, humans can become infected with geohelminths by swallowing infective eggs of the parasite or through the penetration of the skin by infective larvae from the soil.(3) In Nigeria, human and animal wastes are indiscriminately disposed into water and the soil, leading to the contamination of the soil and water with eggs and larvae of these helminths. This increases the risk of people coming in contact with faeces that contain infective eggs and larvae.(4) Almost 2 billion people (a quarter of the world’s population) are infected with geohelminths worldwide, with the greatest numbers reported in Asia, sub-Saharan Africa and the Americas.(5) Nigeria is reported to have the highest number of infected people in sub-Saharan Africa and every state in Nigeria is endemic for geohelminth infections.(6,7) Infection with geohelminths affect virtually all members of a population, but the most vulnerable groups are the pre-school and school age children with severe consequences on their physical and mental health.(8) Children are prone to geohelminth infections because of their less developed immunity, poor toilet habits, tendency to walk bare footed, poor personal hygiene and poor hand washing practices.(9) Geohelminth infections usually result in malnutrition, iron deficiency anaemia, mal-absorption syndrome, intestinal and biliary obstruction, intestinal bleeding, chronic dysentery, rectal prolapse, respiratory complications and impaired cognition in children.(10) All these conditions affect a child’s growth and impede the ability of a child to assimilate properly in class. These can have long term consequences on an individual’s social and professional life. The WHO has recommended some approaches to be able to achieve sustained control of the prevalence, infection intensity and morbidity from geohelminth infections. The approach includes access to appropriate sanitation, hygiene education and preventive chemotherapy.(11) An important measure in controlling geohelminth infections is the use of surveys like this to assess the prevalence of the disease among a given population. There is paucity of information on the prevalence of geohelminth infections among primary school children in Qua’an-Pan Local Government Area, Plateau State. This study seeks to provide baseline information on the diagnoses, planning and implementing of control programmes against geohelminth infections. This study will also add to existing literature on the spread and prevalence of geohelminth infections in Nigeria.

## Materials and Methods

### Study Area and Population

Qua’an-Pan LGA is located in the southern region of Plateau state and is composed of eight districts namely; Bwall, Doemak, Dokan Kasuwa, Kwa, Kwalla, Kwande, Kwang and Namu. Qua’an-Pan LGA is located on latitude 8° 48′ N and longitude 9° 9′ E and covers an area of about 2,478 km^2^.(12) Qua’an-Pan LGA has a tropical climate with average annual temperature of 27.4 ºC. The average humidity level is at 52% and precipitation averages 1,235 mm.(13)

### Sample Size Determination

Sample size for the determination of geohelminth infection was calculated according to the Cochran’s formula (14):

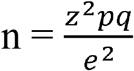

n = sample size

z (standard normal distribution at 95% confidence interval) = 1.96 p (prevalence) = 50% (15)

p = 0.5

q = 1-p = 1 - 0.5

q = 0.5

e = the allowable error, taken as 5% = 0.05

The minimum sample size calculated for this study was 384 children. However, a total of 806 children were included in this study. Random sampling was used to collect data from children within 5 districts in Qua’an-Pan LGA (Kwalla, Bwall, Kwang, Dokan Kasuwa and Kwande).

### Study Design

The cross-sectional study was carried out in schools within each of the 5 selected districts from September, 2021 to December, 2021. A total of 10 primary schools were selected; two schools from each selected district. The two schools were made up of a private and a public school. Samples were collected from at least 43 children per school.

### Ethical Clearance and Consent

Permission was obtained from the Plateau State Ministry of Health (Ref: MOH/MIS/202/VOL.T/X) and Qua’an-Pan Local Government Council (QPLG/S/OFF/VOL.1/114) respectively. Ethical clearance was obtained from the Committee on Use of Human Subjects for Research, Ahmadu Bello University, Zaria (ABUCUHSR/2021/030). Informed consent was obtained through signed consent letters from the parents. Also, oral consent was obtained from the Head-teachers and pupils of each sampled school.

### Inclusion and Exclusion Criteria

Included in this study were; children that were willing to participate, children whose parents/guardians signed the consent forms and children that were able to provide samples on the days of sample collection. In contrast, children with apparent disability and those that were not able to present samples were excluded from the study.

### Sample and Data Collection

The study participants were provided with tissue paper and a labelled sample collection bottle each, they were also given instructions on how to deposit about 2g of stool into the bottles by visible illustrations. Water and detergent were provided for each participant to wash their hands after submitting their samples. About 3 ml of 10% formalin was added to the stool and urine samples that were returned. Formalin acted as a fixative. The stool samples were kept in a box and transported to the laboratory for parasitological examination.

### Examination of Stool Samples

The stool samples were examined at the Helminthology Laboratory of the Department of Parasitology and Entomology, Faculty of Veterinary Medicine, Ahmadu Bello University, Zaria. Each stool sample was examined for eggs and larvae using the Formalin-ether concentration (FEC) technique for geohelminths.(16) About 1g of the faeces mixed with physiological saline was put in a screw-cap bottle containing 4ml of 10% formol water. The bottle was capped and mixed by shaking for about 20 seconds. Thereafter, the faeces were sieved, and the sieved suspension was collected in a beaker. The suspension was transferred to a tube and 3ml of ether was added. The tube was stopped and mixed by shaking for one minute. Thereafter, the stopper was removed and centrifuged immediately at 3000 rpm for one minute. After centrifuging, four layers became evident; the top layer of ether, thin layer of debris, formalin, and sediment in bottom with parasites. An applicator stick was used to loosen the layer of faecal debris from the side of the tube. The ether, debris and formalin were carefully poured off. The sediment was mixed, transferred to a slide and covered with a cover glass. The slide was examined under the microscope using first, the 10x objective followed by 40x objective to identify the eggs. The WHO Bench Aids for the diagnosis of intestinal parasites was used to identify the different geohelminth eggs.(17)

### Data Analyses

SPSS version 20.0 was used to analyze data. Data was displayed in means, frequencies and percentages. Chi-squared test was used to determine the strength of association of potential risk factors with geohelminth infections. P ≤ 0.05 was considered statistically significant.

## Results

### Prevalence of Geohelminth Infections in Qua’an-Pan Local Government Area

Table 1 presents data on the prevalence of geohelminth infections in the five districts that were sampled in Qua’an-Pan LGA. Three geohelminths were identified; they were *Ascaris lumbricoides*, hookworm and *Trichuris trichiura*. The overall prevalence of geohelminth infections was 16.25%. Among all the infected children, hookworm had the highest overall prevalence with 12.40% of the children infected while *Trichuris trichiura* had the least prevalence with 1.61% of the children infected with the parasite. Further analysis with chi-square revealed that there was no significant difference (χ^2^= 3.40, p = 0.18) in the prevalence of each geohelminth infection. Dokan Kasuwa district had the highest overall prevalence (20.0%) of geohelminth infections while Kwalla district had the lowest overall prevalence of geohelminth infections (11.32%) in Qua’an-Pan Local Government Area. Chi-square analysis revealed that there was no statistical difference (χ^2^=0.44, p=0.98) in the prevalence of geohelminth infections among the districts sampled in Qua’an-Pan Local Government Area, Plateau State, Nigeria.

**Table 1:**
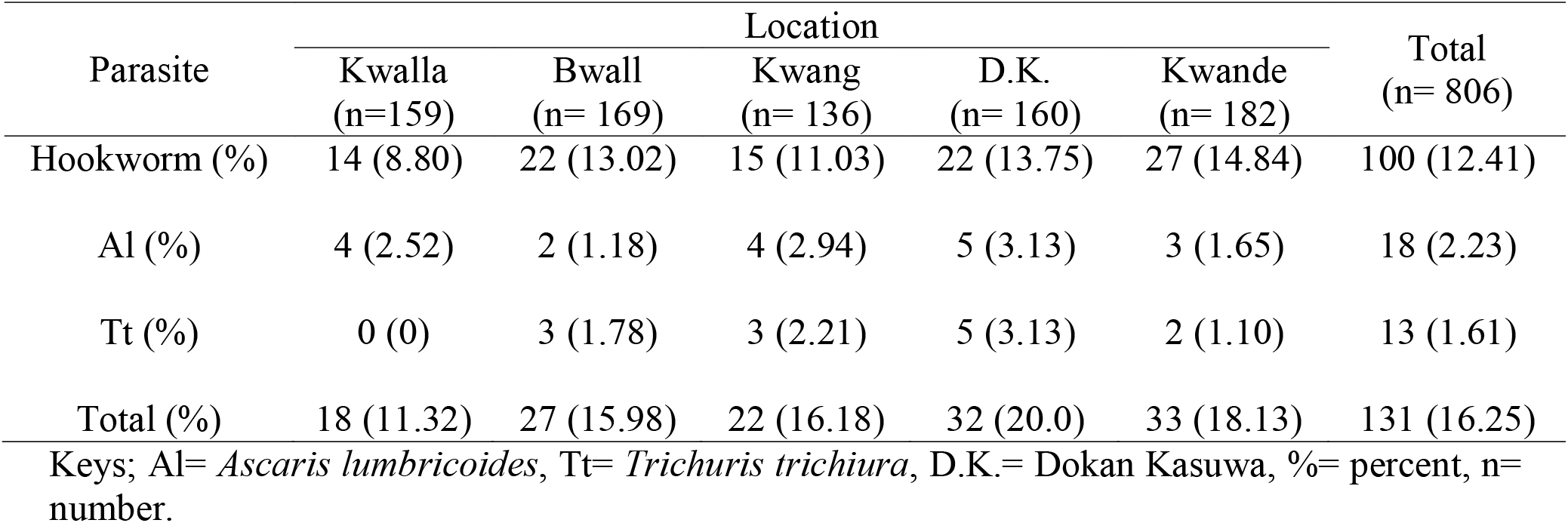
Prevalence of Geohelminth Infections in Qua’an-Pan Local Government Area.

### Prevalence of Geohelminth Infections Based on Sex in Qua’an-Pan Local Government Area

Table 2 shows the prevalence of geohelminth infections based on sex among primary school children in Qua’an-Pan Local Government Area. A total of 409 males and 397 females were examined. The male children had a higher overall prevalence (17.34%) than the female children (14.36%). Chi-square analysis showed that there was no statistical difference in the prevalence of geohelminth infections between the male and female children in Qua’an-Pan Local Government Area (χ^2^= 0.18, p= 0.67).

**Table 2:**
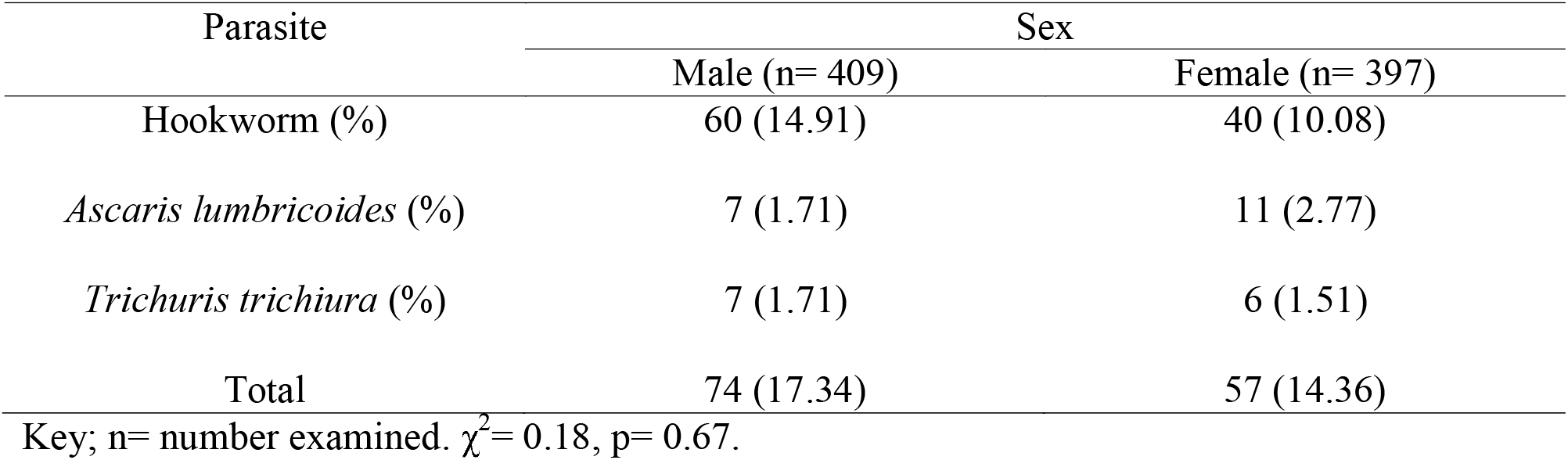
Prevalence of Geohelminth Infections Based on Sex in Qua’an-Pan Local Government Area.

| Parasite | Sex |  |
| --- | --- | --- |
|  | Male (n= 409) | Female (n= 397) |
| Hookworm (%) | 60 (14.91) | 40 (10.08) |
| *Ascaris lumbricoides* (%) | 7 (1.71) | 11 (2.77) |
| *Trichuris trichiura* (%) | 7 (1.71) | 6 (1.51) |
| Total | 74 (17.34) | 57 (14.36) |

Key; n= number examined. $\chi^2 = 0.18$ , $p = 0.67$ .

### Prevalence of Geohelminth Infections Based on Age Groups in Qua’an-Pan Local Government Area

Table 3 shows the prevalence of geohelminth infections among primary school children in Qua’an-Pan Local Government Area based on the following age groups; 5-9 years, 10-14 years and 15-19 years. The 10-14 years group had the highest overall prevalence (17.50%) of geohelminth infections while the children between the ages of 15-19 years had the lowest overall prevalence of geohelminth infections (9.43%). Chi-square analysis showed that there was no statistically significant difference in the distribution of geohelminth infections among the three age groups that were studied (χ^2^= 0.19, p= 0.91).

**Table 3:**
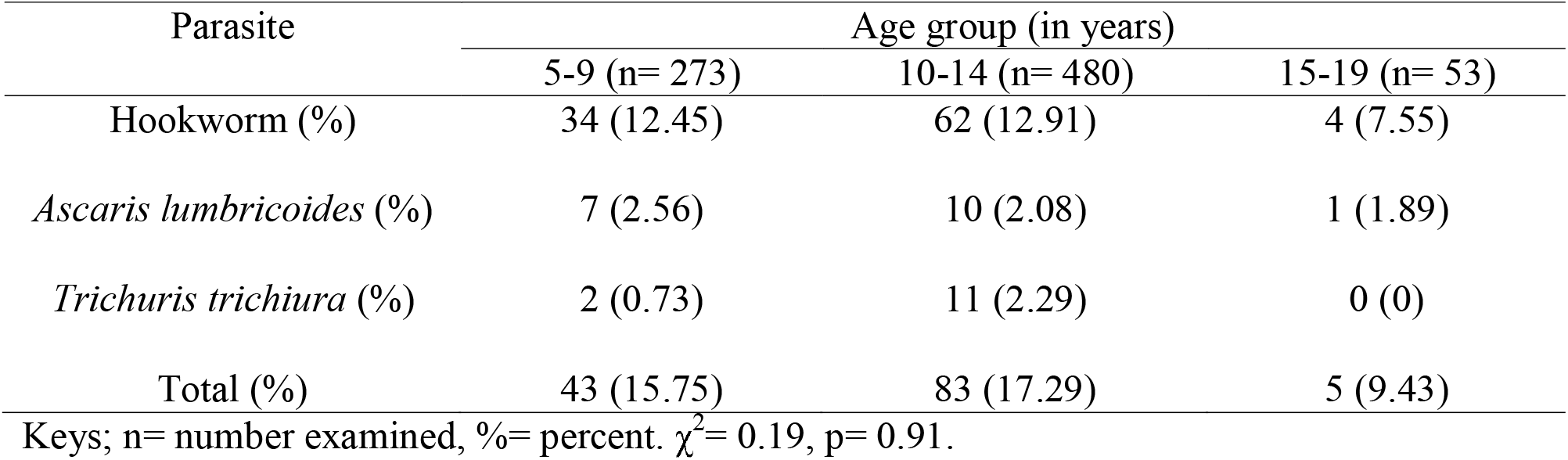
Prevalence of Geohelminth Infections Based on Age Groups in Qua’an-Pan Local Government Area.

### Prevalence of Geohelminth Infections Among Children Attending Public and Private Primary Schools in Qua’an-Pan Local Government Area

Table 4 displays the prevalence of geohelminth infections in the public and private schools. Overall, the public schools had a higher prevalence (17.26%) than that of the private schools (15.17%). Chi-square analysis was used to show that there was no statistical difference in the overall prevalence of geohelminth infections between the public and private schools (χ^2^= 0.06, p= 0.81).

**Table 4:** Prevalence of Geohelminth Infections Among Children Attending Public and Private Schools in Qua’an-Pan Local Government Area.

| Parasite | Type of School attended |  |
| --- | --- | --- |
|  | Public (n= 417) | Private (n= 389) |
| Hookworm (%) | 55 (13.19) | 45 (11.57) |
| *Ascaris lumbricoides* (%) | 9 (2.16) | 9 (2.31) |
| *Trichuris trichiura* (%) | 8 (1.92) | 5 (1.29) |
| Total (%) | 72 (17.27) | 59 (15.17) |

Keys; n= number examined, %= percent. $\chi^2 = 0.06$ , $p = 0.81$ .

## Discussion

The overall prevalence of geohelminthiasis in this study was 16.25%. This figure is below the 20% baseline mark set by the WHO for annual distribution of single dose anti-helminthic drugs. This means that there is no immediate need for the administration of anti-helminthic drugs yet but it is a situation that needs to be closely monitored. In comparison with similar studies, the prevalence obtained from this study is lower than the prevalence reported in Jos North Local Government Area (24.3%) and Jos South Local Government Area (42.6%) in Plateau State, Nigeria.(18,19) A reason for the relatively low prevalence of geohelminth infections in Qua’an-Pan Local Government Area could be the periodical administration of preventive chemotherapy against schistosomiasis and geohelminth infections among schools found in the study area (mostly the public primary schools). This was confirmed on interaction with some Head-teachers of the schools that were sampled. The Head-teachers pointed out that the distribution of the therapeutic drugs was not regular. It was also observed that the majority of the sampled schools didn’t have toilets and, in the case, that toilets were present, they were inadequate and not well kept. School children were allowed to ease themselves in the farms that were close to their school. This habit is known to ensure the maintenance and transmission of parasites among school children.

Hookworm infections were more prevalent than any other geohelminth infection in Qua’an-Pan Local Government Area. This is similar to the findings in Jos North Local Government Area and in Jos South Local Government Area, both situated in Plateau State, Nigeria.(18,20) Houmsou and colleagues suggested that the presence of sandy soil, optimum temperature (7°C - 31ºC) and rainfall was essential in sustaining hookworm infections in Benue State. These climatic conditions are known to aid the development the larval stages of hookworms.(21) The presence of similar conditions in Qua’an-Pan Local Government Area can be said to be a reason for the higher prevalence of hookworm infections in this study. Also, walking, playing and working barefoot are habits that allow the infection to be transmitted easily. Children are known to take off their footwear before playing mainly because they want it to last longer. This habit increases the risk of hookworm larvae penetrating the skin of unsuspecting children. Although Kwande district had the highest overall prevalence of geohelminth infections, there was no significant difference between it and the other four districts that were sampled in this study. This implies that the distribution of the infections across the districts were similar. Hence, location was not a risk factor for the infections.

The male children in this study had a higher overall prevalence of geohelminth infections than the female children. This agrees with the work of Houmsou *et al*.(21) A reason for this higher prevalence of geohelminth infections among boys could be because the boys are more likely to engage in risky behaviours such as playing football barefoot and swimming. Also, children within between the ages of 10-14 years and 5-9 years had higher overall prevalence of geohelminth infections than children between the ages of 15-19 years. This finding is similar to the work of Mulumbalah and Ruto.(22) This could be because the younger children are old enough to play unsupervised and are not yet at an age where they begin to take their personal hygiene and appearance seriously. At such an age, play is seen as a priority.

An interesting finding from the study shows that children attending private primary schools in Qua’an-Pan Local Government Area had a higher but statistically insignificant overall prevalence of geohelminth infections than their counterparts that attended public primary schools. This disagrees with the work of Ogwurike and colleagues.(18) It is expected that children that attend private primary schools generally have benefactors that are of a higher socio-economic status than children who attend public primary schools. It is logical to accept this reasoning as access to medical care, attending private schools, good nutrition and enlightenment can be positively associated with a high socio-economic status. A reason for this contrasting finding could lie far away from the schools these children attend. It could be found in the play grounds these children share after school hours. There is also the possibility that they all go swimming in the same water body or obtain drinking water from the same source. It can serve to even the spread of geohelminth infections between the children that attend public and private primary schools in Qua’an-Pan Local Government Area.

## Conclusion

Geohelminth infections are prevalent in Qua’an-Pan Local Government Area of Plateau State, Nigeria. The sex, age group and the type of school attended by the children in the study area did not directly affect the distribution of geohelminth infections. It is advised that government should ensure the annual distribution of antihelminthics to children in Qua’an-Pan Local Government Area and other rural areas to curb the menace of geohelminth infections. Also, enlightenment programmes can be staged while distributing antihelminthic drugs to help children and caregivers know more about these parasitic diseases and how to prevent them. There should also be efforts to ensure that people in rural areas have access to clean water for drinking.

## Data Availability

All data produced in the study are available upon reasonable request to the authors.

## Acknowledgements

Our sincere appreciation goes to the children, parents/guardians, teachers and Head-teachers that participated in this study. Also Mr. Wilberforce Sogotdiel sacrificed a lot to make sure that the researchers had an easy field work. We are most grateful.

